# Effects of Public Health Interventions on the Epidemiological Spread During the First Wave of the COVID-19 Outbreak in Thailand

**DOI:** 10.1101/2020.09.01.20182873

**Authors:** Sipat Triukose, Sirin Nitinawarat, Ponlapat Satian, Anupap Somboonsavatdee, Ponlachart Chotikarn, Thunchanok Thammasanya, Nasamon Wanlapakorn, Natthinee Sudhinaraset, Pitakpol Boonyamalik, Bancha Kakhong, Yong Poovorawan

## Abstract

A novel infectious respiratory disease was recognized in Wuhan (Hubei Province, China) in December 2019. In February 2020, the disease was named “coronavirus disease 2019” (COVID-19). COVID-19 became a pandemic in March 2020, and, since then, different countries have implemented a broad spectrum of policies. Thailand is considered to be among the top countries in handling its first wave of the outbreak -- 12 January to 31 July 2020. Here, we illustrate how Thailand tackled the COVID-19 outbreak, particularly the effects of public health interventions on the epidemiologic spread. This study shows how the available data from the outbreak can be analyzed and visualized to quantify the severity of the outbreak, the effectiveness of the interventions, and the level of risk of allowed activities during an easing of a “lockdown.” This study shows how a well-organized governmental apparatus can overcome the havoc caused by a pandemic.

## Introduction

A novel coronavirus disease is officially recognized in Wuhan, China, in December 2019 [1]. In February 2020, the disease is later named Coronavirus disease 2019 (COVID-19), which is an emerging infectious disease caused by the severe acute respiratory syndrome coronavirus 2 (SARS-CoV-2) [2]. After its first discovery, it had then swiftly spread globally. The World Health Organization (WHO) declared the outbreak of a Public Health Emergency of International Concern on 30 January 2020, and a pandemic on 11 March 2020 [3]. The severity of the outbreak across different countries varies significantly due to several factors, such as timeliness and strength of state interventions, country healthcare readiness, and socioeconomic considerations [4]. In this regard, Thailand has been widely praised for its handling of the COVID-19 outbreak. In particular, it is ranked second for the Global COVID-19 Index (GCI) and first in Asia as of 31 July 2020 [5]. The index is developed by PEMADU Associates, in collaboration with the Ministry of Science and Innovations of Malaysia and the Sunway Group. The WHO has also chosen Thailand and New Zealand to be featured in their upcoming documentary as exemplary countries that have handled COVID-19 most successfully [6].

Since the initial report of cases in Wuhan city on 31 December 2019, the Ministry of Public Health Thailand implemented measures for screening travelers from Wuhan city on 3 January 2020 at Suvarnabhumi Airport, Don Mueang, Phuket, and Chiang Mai airports by checking their body temperature and respiratory symptoms. Enhanced surveillance at public and private hospitals was also initiated. Thailand identified the first case on 12 January 2020 (officially announced on 13 January 2020) [7]. The case was a 61-year-old Chinese woman living in Wuhan City, Hubei Province, China. On 5 January 2020, she developed fever with chills, sore throat, and headache. On 8 January 2020, she took a direct flight to Bangkok from Wuhan City. The febrile illness was detected on the same day by thermal surveillance at Suvarnabhumi Airport, Bangkok, Thailand. She was transferred to the hospital for further investigations and treatment. Clinical samples were tested positive for coronaviruses by reverse transcriptase-polymerase chain reaction (RT-PCR) on 12 January 2020. The genomic sequencing analysis confirmed that the patient was infected with the novel coronavirus (2019-nCoV) [8]. The number of confirmed cases was still low throughout January and February 2020, during which the confirmed cases were mostly from travelers who came from China or other countries [8].

In early March 2020, the number of confirmed cases from local transmission started to increase rapidly. Several transmission clusters contributed to the increased number of confirmed cases in Thailand, the largest of which was at the entertainment venue and Thai boxing stadium in Bangkok in early of March 2020 [8,9].

In response to the escalating situation, on 12 March 2020, the Thai Government established the Center for COVID-19 Situation Administration (CCSA) as a single command center to ensure a coherent view of the situation and unambiguous communication to the public about all related matters.

On 18 March 2020, Thailand medical council declared a concerning statement and demonstrated the first Thai statistical epidemiological model forecasting large outbreak scenarios and their loads on the national healthcare capacity [8]. Since then, the Thai Government has officially implemented multiple disease-controlling and public health policies in response to the COVID-19 situation in Thailand [8].

On 3 May 2020, after a week of a low number of daily confirmed cases, the CCSA announced that Thailand was entering the Easing period and started rolling out policies to relax restrictions and interventions implemented earlier.

On 31 July 2020, when the global cumulative COVID-19 infected and death cases were 1,710,6007 and 668,910 respectively [10], Thailand exited the first wave of COVID-19 outbreak gracefully with the cumulative COVID-19 infected, and death cases being 3,310 and 58, respectively. This success was a collaborative effort of all healthcare-related personnel and all Thais. On the same day, Thailand’s confirmed case was ranked 107 of 213 countries affected by COVID-19 [11].

In this study, the course of COVID-19 outbreak in Thailand is to be described and studied together with the timeline of the Thai Government’s disease-controlling and public health policies that are needed to prepare for the outbreak scenarios. The objectives of this study are the following: i) to report about Thailand’s public health interventions and the epidemiological dynamics of COVID-19 therein during the first wave of the epidemic, and ii) to gather lessons learned from the first wave. In particular, our approach and results are reminiscent of those reported in [1] for Wuhan, China. Still, the contexts of that article and ours are different as the nature of the outbreak (started endogenously and exogenously respectively), interventions, and enforcement in the various cities in two different countries. One of our significant findings is the power of the effective reproduction number to forecast the future course of the epidemic, thereby underlying its importance as a monitoring index that the Government could use to increase the intervention strength at the right time to mitigate the potential surging of the epidemic. Given how well Thailand has handled the first wave, we hope that the learned lessons are useful for other countries, the general public, and Thailand itself during the second wave if it were to occur.

## Materials and methods

### Data sources

This study used the medical records of laboratory-confirmed COVID-19 patients in Thailand from 12 January 2020 to 31 July 2020. These medical records were retrieved from the Department of Disease Control of Thailand website [12]. The retrieved data set comprises age, sex, nationality, date of confirmed COVID-19-positive, location of onset, isolation, and quarantine history. The information about public health policies and critical events in this study was extracted from the CCSA, and Thai Government official reports.

### Definition of the first-wave period

We defined the first wave of COVID-19 outbreak in Thailand as the period from 12 January 2020, when the first imported case was identified (officially confirmed on 13 January) to 31 July 2020, one month from the beginning of the 5th easing period (see the easing periods section) and the point at which Thailand had no local infectious report for 67 consecutive days.

### Classification of the five time periods

In reflecting the dynamics of the first wave COVID-19 epidemic and its relationship to corresponding interventions in Thailand, the first-wave period was classified into five epidemic stages (Fig 1): (A) Early; (B) Spreading; (C) Intervention I; (D) Intervention II; (E) Easing. This classification was based on critical events as well as public health interventions and policies (Fig 2).

**Fig 1.**
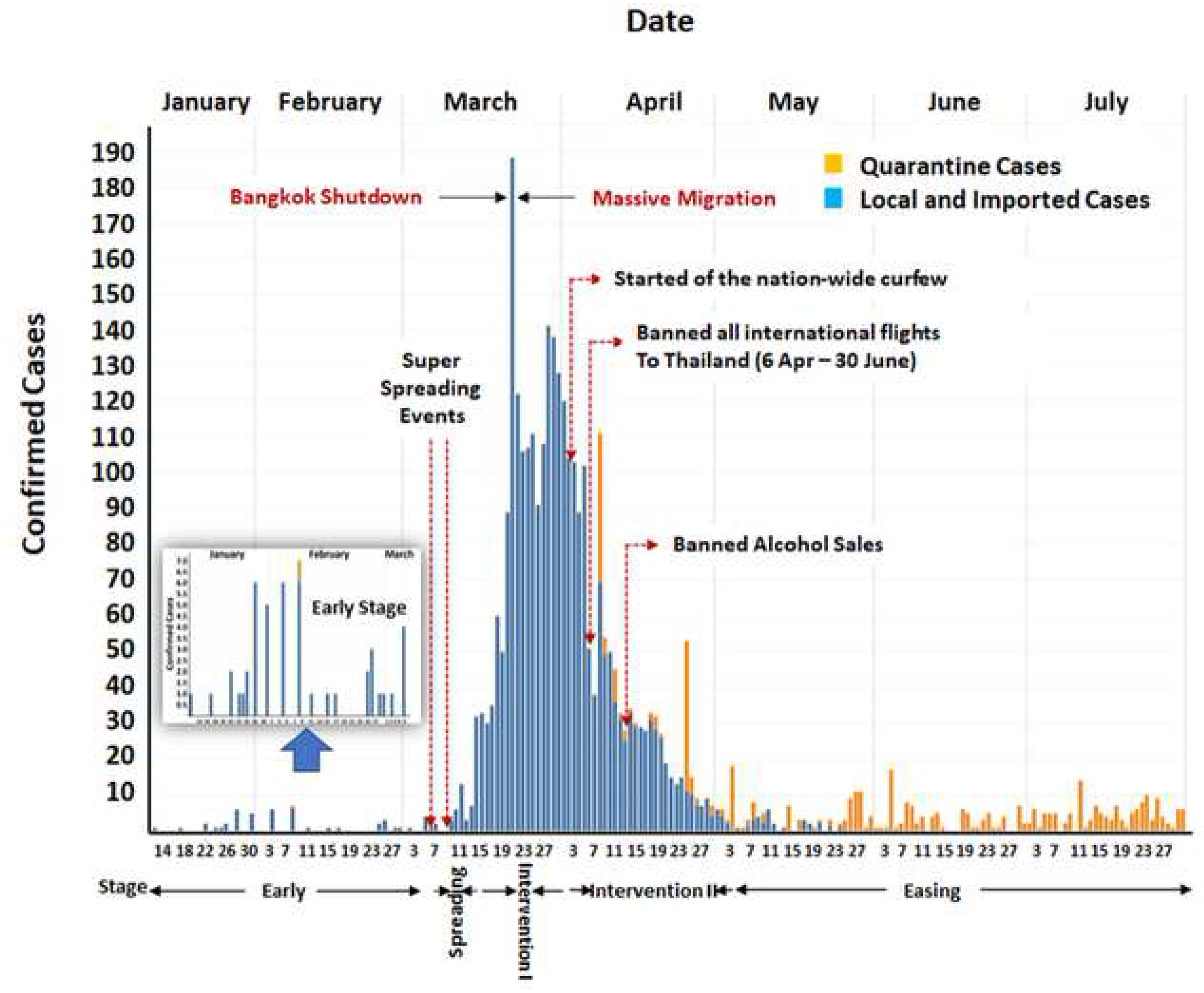
Epidemic curve across five stages during the COVID-19 outbreak in Thailand.

**Fig 2.**
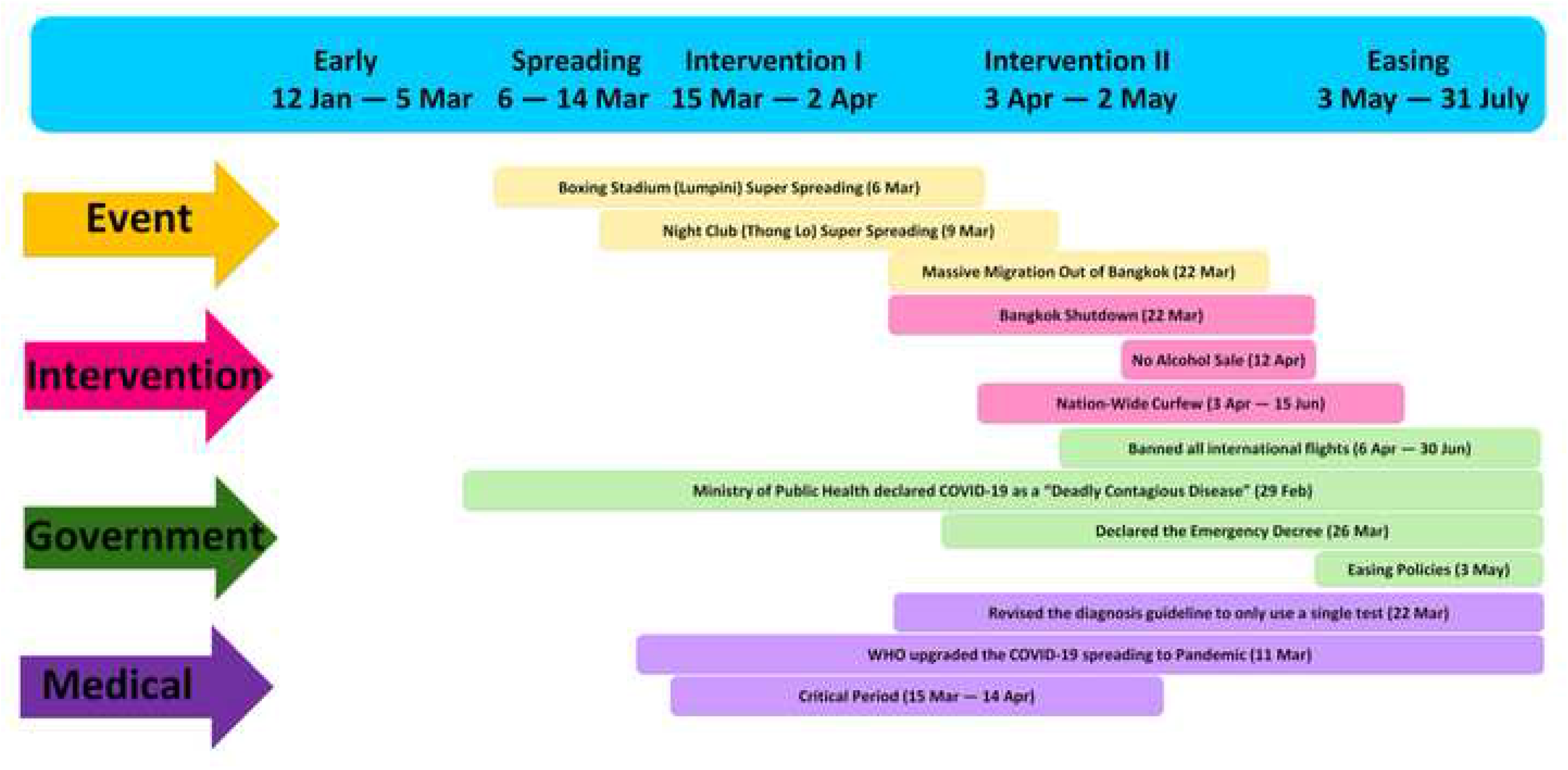
Timeline of key events and public health interventions across the five stages of the COVID-19 epidemic in Thailand.

The Early stage (A) started from 12 January to 5 March 2020. The first imported and local cases were identified on 12 January and 31 January 2020, respectively. On 1 March 2020, Thailand had the first patient who died from COVID-19. During this stage, the total number of COVID-19 cases was below 100 cases, and there was no strong public health intervention. However, Thais were quite active in wearing facial masks due to the PM2.5 crisis during that time.

The Spreading stage (B) was from 6–14 March 2020. This stage was a short period but contained two “super-spreading” events in Bangkok: at the Lumpini boxing stadium on 6 March 2020 and an entertainment venue in the Thong Lo area on 9 March 2020. These events contributed to the outbreak in Thailand [9]. After this stage, the number of total COVID-19 confirmed cases was higher than 100 cases, and Thailand entered the critical period of infectious disease. The critical period (from 15 March to 14 April 2020 in the case of Thailand) is defined as a 30-day duration from the first time at which the number of total confirmed cases is higher than 100 cases [13]. It is believed to be an extremely crucial moment in combating an epidemic because actions during this period dictate if the outbreak will be in or out of control.

The Intervention I stage (C), and Intervention II stage (D) were the stages that all Thais put the people’s lives in front of everything. Several public health intervention policies were implemented, and most people cooperated. The Intervention I stage started at the same time at the beginning of the critical period and lasted until 2 April 2020. During this period, local and governmental authorities focused on reducing and preventing all social-gathering activities. The Lumpini boxing stadium and the entertainment venue in Thong Lo were closed down on 15 March 2020. The Songkran festival (Thai New Year) -- a long holiday (similar to Christmas) in which numerous people return home to reunite with their parents and loved ones -- was canceled on 16 March 2020. Several public venues were closed down on 18 March 2020. Finally, on 22 March 2020, the Bangkok mayor shut down the city, resulting in the suspension of many jobs. Unfortunately, the leaked news of the shutdown led to a massive migration of workforces in Bangkok back to their hometown just before the actual shutdown. This was the key event that caused the outbreak to spread countrywide. Therefore, the Intervention II stage, from 3 April to 2 May 2020, involved additional policies to control the route of transmission. A nationwide curfew was implemented on 2 April 2020, and the Civil Aviation Authority of Thailand (CAAT) banned all international flights to Thailand starting from 6 April 2020. A mandatory state quarantine was established on 3 April 2020, for everyone traveling to Thailand. Alcohol sales were prohibited from 12 April 2020, throughout the Intervention II period [9].

At the end of the critical period, public health intervention policies that local and Government executed during the Intervention I and Intervention II stages were successful: the number of daily confirmed cases in Thailand returned to the same level before the critical period. Therefore, Thailand entered the Easing stage (E) from 3 May to 31 July 2020. In this stage, restrictions correlated to the lower infectious risk were lifted and observed for 14 days before proceeding to the next level, which eased the policies with higher contagious risk. In addition, we compared the trade-off of the two different COVID-19 handling approaches on health and economics by selecting two neighbours in Scandinavian, Sweden, Denmark, and Thailand as subjects of our analysis. Sweden has been one of the countries that adopt the herd immunity strategy. In contrast, Denmark and Thailand have used lockdown and social distancing measures to limit local cases.

### Outcomes

The number of daily confirmed cases is the total new laboratory-confirmed cases on a particular day. It is interesting to note that the Government initially used a stringent criterion for the case to be confirmed, requiring each individual to be approved by the two assigned national laboratories. Later, on 22 March 2020, the criterion was changed, and a new case was confirmed using just one national laboratory result, instead of two [14].

### Statistical analysis and data visualization

Unless specified otherwise, all the data clean up, transformation, and calculation were done using R and Python languages. The epidemic curves were generated by Tableau software. Tables and Annotations on Figures were done using Apple Keynote and Microsoft Excel.

We follow the general statistical framework by Cori *et. al*. [15] to estimate the effective reproduction number at day *t* (*R*_t_) and its credible band of all epidemic stages [15]. Under this framework, the number of infected cases at day *t (I_t_)* is assumed to have a Poisson distribution with the rate of *R_t_* (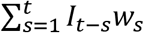) where *w_s_*’s are weights derived from the serial interval distribution of the disease. Moreover, in this study, the serial interval distribution was assumed to be gamma distribution with a mean and standard deviation of 7.5 and 3.4 days, respectively, as described by Pan *et al*. [1], where they also applied the framework precisely to the COVID-19 epidemic in Wuhan, China. Under the Bayesian framework, the gamma distribution is used as a prior distribution for the effective reproduction number where the obtained posterior distribution will be used to estimate the effective reproduction number at time *t* (*R*_t_) and its credible band (see the supplementary of Cori *et al*. [15] for more detail).

We applied the mentioned methodology on the data from Thailand (country-level) and the two hotspot provinces: Bangkok (the capital city) and Phuket. The estimates of the effective reproduction numbers and their 95% credible bands and daily laboratory-confirmed cases in the respective regions are shown.

The Thailand geographic spread maps were created using the Quantum Geographic Information System (QGIS). The data were separated into five epidemic stages (A-E). After that, each dataset was plotted geographically using QGIS.

The state and local quarantine cases from the daily confirmed cases were excluded when producing the reproduction number, the Thailand demographic maps, and the characteristics of daily new confirmed COVID-19 cases to highlight local transmission of the disease. The state and local quarantine cases only appear only in the epidemic curves.

## Results and Discussion

### Data characteristics

The characteristics of new daily confirmed COVID-19 cases across the five epidemic stages in Thailand are shown in Table 1. The average daily number of confirmed cases was highest during the Intervention I stage (1739). In the Intervention II stage, the average daily number of confirmed cases declined to 967. Table 1 shows that most of the cases were in the age groups 20-29 years and 30-39 years, and there was no apparent association with sex.

**Table 1.** Characteristics of daily new confirmed COVID-19 cases across the five epidemic stages in Thailand.

| Characteristics | COVID-19 epidemic stages |  |  |  |  | Total |
| --- | --- | --- | --- | --- | --- | --- |
|  | Early | Spreading | Intervention I | Intervention II | Easing |  |
|  | 12 Jan 2020 - 5 Mar 2020 | 6 Mar 2020 - 14 Mar 2020 | 15 Mar 2020 - 2 Apr 2020 | 3 Apr 2020 - 2 May 2020 | 3 May 2020 - 31 July 2020 |  |
| **Total** | 46 | 35 | 1793 | 967 | 31 | 2872 |
| **Sex (%)** |  |  |  |  |  |  |
| **Male** | 26 (57) | 17 (49) | 1024 (57) | 442 (46) | 17 (55) | 1526 (53) |
| **Female** | 20 (43) | 18 (51) | 769 (43) | 525 (54) | 14 (45) | 1346 (47) |
| **Age group (%)** |  |  |  |  |  |  |
| **0-19** | 3 (7) | 2 (6) | 69 (4) | 74 (8) | 3 (10) | 151 (5) |
| **20-29** | 9 (20) | 8 (23) | 488 (27) | 227 (23) | 7 (23) | 739 (26) |
| **30-39** | 12 (26) | 15 (43) | 451 (25) | 226 (23) | 4 (13) | 708 (25) |
| **40-49** | 4 (9) | 8 (23) | 332 (19) | 183 (19) | 9 (29) | 536 (19) |
| **50-59** | 5 (11) | 1 (3) | 249 (14) | 140 (14) | 6 (19) | 401 (14) |
| **60-69** | 9 (10) | 1 (3) | 132 (7) | 72 (7) | 0 (0) | 214 (7) |
| **70-79** | 4 (9) | 0 (0) | 51 (3) | 26 (3) | 1 (3) | 82 (3) |
| **≥ 80** | 0 (0) | 0 (0) | 11 (1) | 12 (1) | 1 (3) | 24 (1) |
| **N/A** |  |  | 10 (1) | 7 (1) |  | 17 (1) |
| **Tested population** | N/A | N/A | N/A | 149875 | 381747 | 531622 |

### Geographic spread and new confirmed cases

This section reports Thailand’s geographic information for the average daily number of confirmed cases. Fig 3 shows how COVID-19 spread across the country in five epidemic stages. In the Early stage, the COVID-19 was mostly limited to Bangkok (Fig 3A), in which the average daily number of confirmed cases was less than 1.

**Fig 3.**
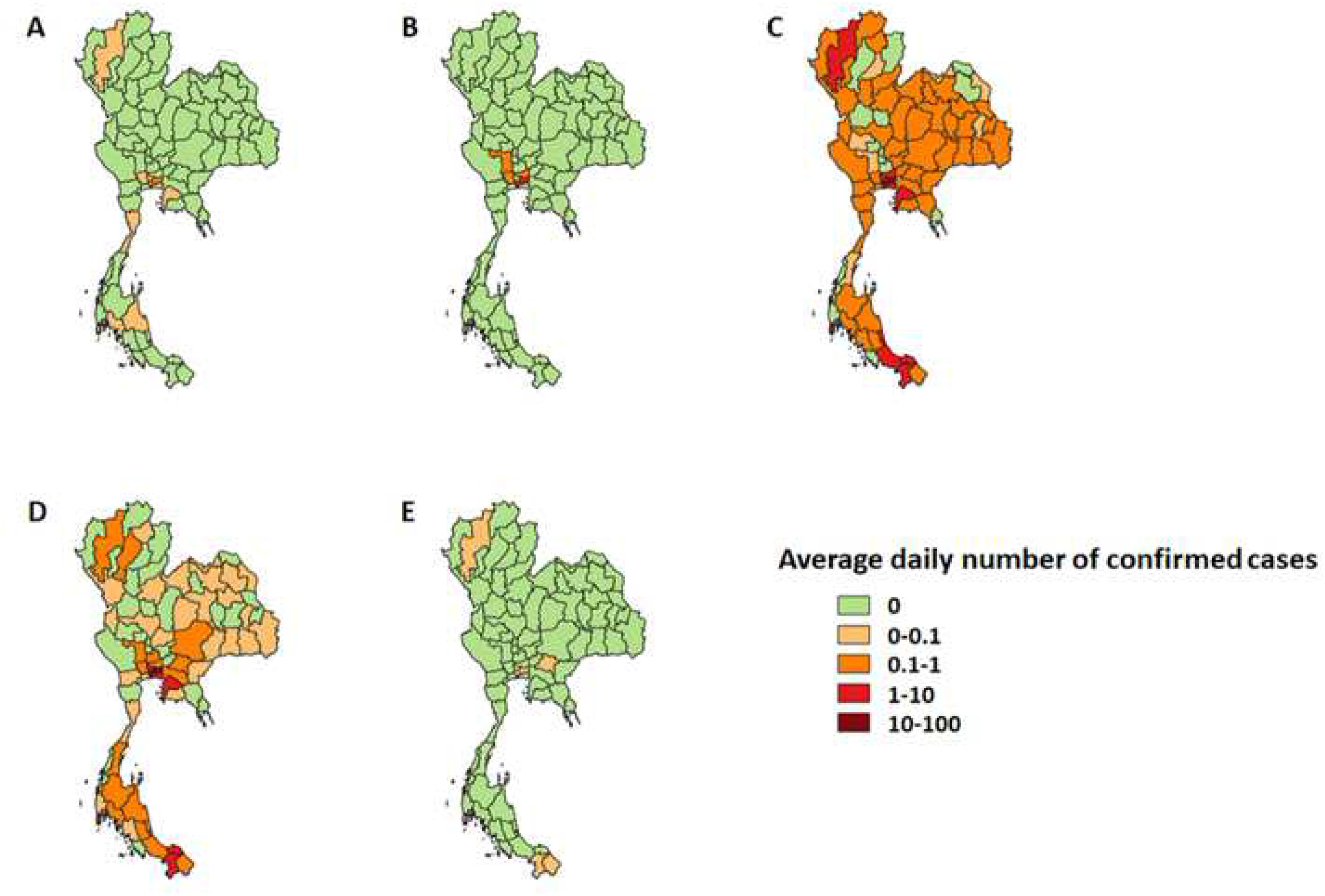
Geographic spread of the average daily number of confirmed cases in the five epidemic stages of COVID-19 in Thailand. (A) Early stage (B) Spreading stage (C) Intervention I stage (D) Intervention II stage (E) Easing stage

SARS-CoV-2 started to spread across Bangkok’s border during the Spreading stage (Fig 3B). The highest average daily number of confirmed cases, found in Bangkok, was less than 10.

In the Intervention I stage (Fig 3C), COVID-19 swiftly spread nationwide mainly due to a massive migration back home after the Bangkok shutdown on 22 March 2020. During this stage, the highest average daily number of confirmed cases, found in Bangkok, was 54.83. The average daily number of confirmed cases in Nonthaburi, Phuket, Samutprakan, Chonburi, Pattani, Yala, Chiangmai, Songkhla, and Pathumthani was 5.67, 4.78, 4.44, 2.56, 2.56, 2.22, 1.89, 1.39 and 1.22 cases per day, respectively. In contrast, only 14 of 77 provinces had no cases in this epidemic stage.

In the Intervention II stage, the public health interventions seemed to be successful, and the infectious rate declined and became stable (Fig 3D). Bangkok had the highest average daily number of confirmed cases (14.79). Only six provinces (Phuket, Yala, Nonthaburi, Chonburi, Samutprakan, and Pattani) had the average daily number of confirmed cases between 1 to 10.

Lastly, in the Easing stage, the average daily number of confirmed cases dropped to zero almost nationwide (Fig 3E). The new confirmed cases came from nine provinces: Ang Thong, Bungkan, Chainat, Kamphaengphet, Nan, Pichit, Ranong, Singburi, and Trat.

### Effective reproduction number

The effective reproduction number or *R_t_* is the expected number of secondary subjects infected by a primary subject at day *t*. It is commonly used to measure the transmission level of infectious disease [15]. In this study, using the framework mentioned in the earlier section, the available data on the confirmed cases at country-level and selected province-level were used to estimate the effective reproduction numbers (*R_t_*’s) and their 95% credible bands, as shown by the red curves and bands, respectively in Fig 4. For the province-level results, we selected two hotspot provinces for the outbreak: Bangkok and Phuket. (Real-time results for Thailand and all available provinces can be found on Thailand COVID-19 *R_t_* Tracker website: https://thai-covid19.live.) It is important to note that the limitations of the data used in the estimation reflected the reliability of the results. The main limitations were (i) a delay in time at which the cases were recorded and confirmed from the actual onset time of disease; (ii) a considerably small number of confirmed cases, especially at the province level.

**Fig 4.**
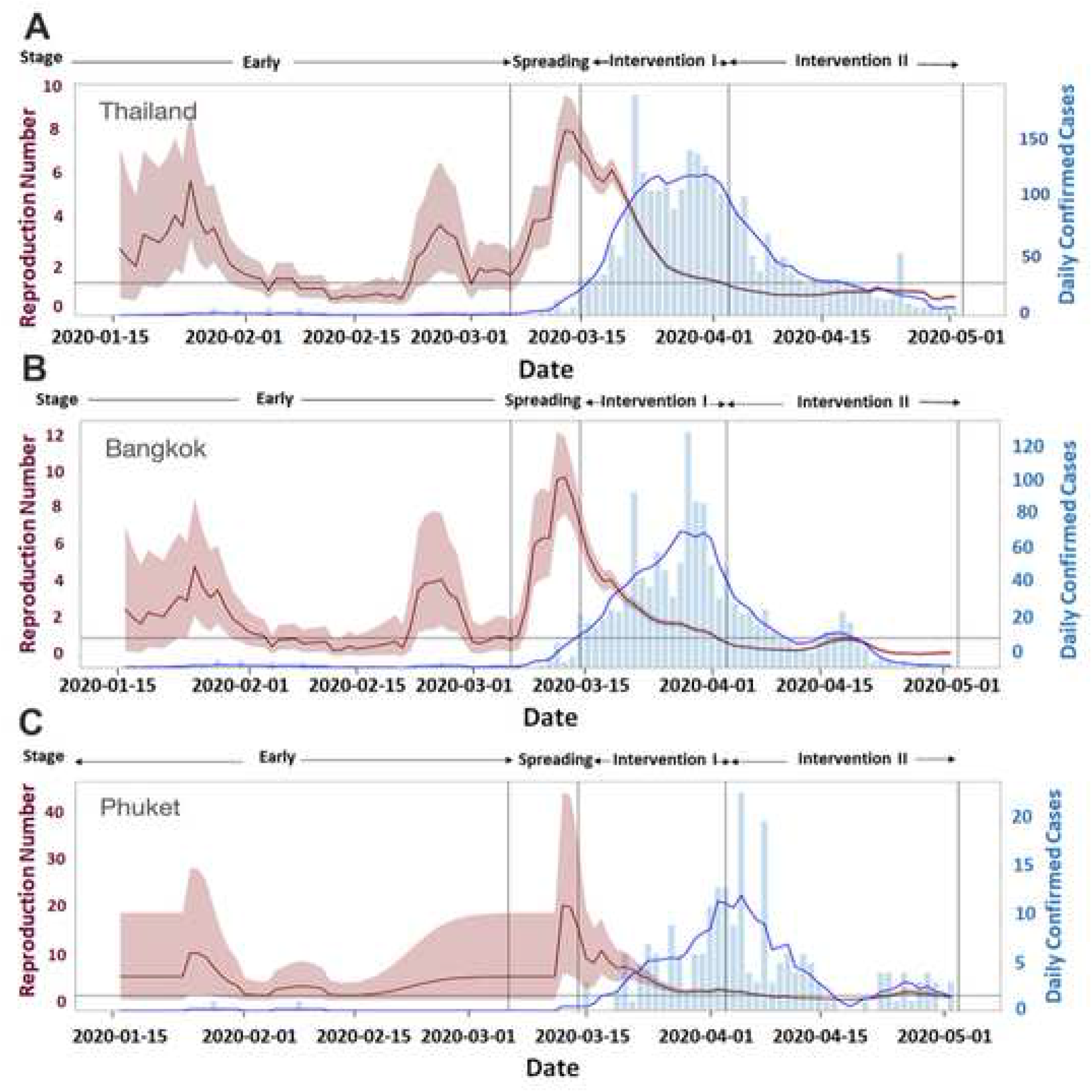
The Effective Reproduction Number *(R_t_)* for COVID-19 Outbreak in Thailand (4A), Bangkok (4B), and Phuket (4C).

The results from Thailand (Fig 4A), Bangkok (Fig 4B), and Phuket (Fig 4C) were similar in overall trends and patterns. More specifically, the results for Thailand and Bangkok were almost identical because most of the confirmed cases in Thailand were from Bangkok, particularly in the Early and Spreading stages. On the other hand, the *R_t_* plot for Phuket in Fig 4C seems to be at a higher level with a broader credible band compared to Thailand’s and Bangkok’s *R_t_* plot. Due to a considerably smaller number of confirmed cases in Phuket, the estimated effective reproductive numbers and its credible band may be less reliable and more conservative than those for Thailand and Bangkok.

In the Early stage, the *R_t_* plots for Thailand, Bangkok, and Phuket showed instability (somewhat randomly up and down) with the daily confirmed cases less than 5 cases per day. These unstable or random trends with very few confirmed cases per day were due to the fact that this was the Early stage when people were starting to become infected.

In the Spreading stage, all *R_t_* plots in Fig 4 show some steep increasing trends, especially for Thailand and Bangkok. There was a bit of a delay in the increasing trend in the *R_t_* plot for Phuket. Once again, these increasing trends of *R_t_* plots collaborated the fact that this period was the Spreading stage of the disease, in which there was a massive increase in the number of infected cases.

In the Intervention I and II stages, all *R_t_* plots showed decreasing trends, supporting the notion that the interventions were effective in preventing the transmission of the disease. More specifically, the *R_t_* plots in the Intervention I stage decreased faster than those in the Intervention II stage. This observation is consistent with the objective of the Intervention I stage, which is to stop and contain the outbreak as fast as possible, whereas the objective of the Intervention II stage was only to keep the outbreak under control.

It can be seen that we can use the trend of the *R_t_* plot and the number of daily confirmed cases in order to identify the stages of the disease outbreak. With more accurate and contemporary real-time data, *R_t_* plot can be used as a monitoring and policy-decision-making tool.

### The Thai Health care system for COVID-19 patients

The clinical criteria for suspicious cases of COVID-19 in Thailand as of 27 February 2020 were: body temperature higher than 37.5°C; cough; rhinorrhea; sore throat; dyspnea or difficulty in breathing; pneumonia of undetermined cause or a cluster of acute respiratory tract infections of undetermined cause. Anosmia was appended to the list of suspect symptoms on 1 May 2020.

The clinical criteria were considered in conjunction with epidemiologic criteria: (i) association with the active areas of COVID-19 transmission (a history of traveling to the areas, or a family member returning from the areas); (ii) in close contact with international travelers or anyone (especially healthcare-related personnel) who was in close contact with a confirmed case within 14 days before symptom onset.

People suspected of having COVID-19 were tested to find SARS-CoV-2 RNA in their clinical specimens. Asymptomatic or symptomatic close contact with a confirmed case of COVID-19 was asked to visit a hospital for a nasopharyngeal-swab test for SARS-CoV-2 RNA. It was mandatory for all SARS-CoV-2 positive patients, regardless of their symptoms, to stay in hospital. People who were asymptomatic or who had mild symptoms were admitted and usually spent 2–7 days in a single isolation room or a cohort ward. Confirmed cases with mild symptoms and comorbidities, or confirmed cases with pneumonia, could not be discharged before full recovery and SARS-CoV-2 negative.

Following hospital discharge, they were mandated to stay in a designated hostel, namely “hospitel,” until 14 days after symptom onset or when their swab results were SARS-CoV-2 negative at least two consecutive times.

Village Health Volunteer (VHVs) is well-established and became one of the essential factors to help control the COVID-19 outbreak in Thailand. Thai Government manages 1,040,000 Village Health Volunteers (VHVs) across the country plus 15,000 public health volunteers in Bangkok.

After receiving training, each volunteer looks after 10-15 households, often home to the bedridden, the disabled, and the elderly. VHVs have spread out across the country to promote public health education, deliver medicines to Noncommunicable diseases (NCDs) patients so that they can stay at home, and make reports to public health authorities.

In light of the COVID-19 outbreak, VHVs have been a key instrument for promoting related public health policies and providing essential supplies, such as facial masks, face shields, biohazard bags, and alcohol gel. Contributing significantly to the outbreak control, VHVs visited more than 11 million households (3.3 million households from 2 March 2020 to 26 March 2020, and 8 million households from 27 March 2020 to 11 April 2020) to help facilitate case finding efforts.

We believe that these measures, imposed for every patient, and VHVs’ network contributed greatly to the excellent outcome of handling the COVID-19 outbreak in Thailand.

### Effect of Public Health Interventions and Key Events

The super-spreading clusters at the Lumpini boxing stadium on 6 March 2020 and the entertainment venue in Thong Lo on 9 March 2020 were the two most important contributors to spreading COVID-19 in Thailand. The pinnacle of this outbreak, 188 confirmed cases on 22 March 2020, was approximately 14 days after the two super-spreading events.

In handling the outbreak, two critical executive decisions played a major part during the Intervention I stage to get Thailand past the outbreak peak before the end of the critical period.

At the end of the critical period (14 April 2020), the number of daily confirmed cases was 33, whereas 32 cases were at the beginning.

The first important decision was enforcement of a set of policies to reduce and prohibit social gatherings. From 15 to 22 March, the Thai Government and Local Governments canceled the Thai New Year (Songkran Festival), in which people gather, roam around, and splash water on each other. The Lumpini boxing stadium, cinemas, sports clubs/complexes, department stores, seated restaurants, and most public-gathering places were closed. The crucial decisions were to: establish an emergency-response mechanism by declaring the COVID-19 as a dangerous communicable disease under the Communicable Diseases Act; declaring the state of emergency; creating the CCSA to respond to the situation promptly; making all communications coherent; gathering all expert advice; fighting the outbreak based on a holistic view of the situation.

A couple of surges on the number of daily confirmed cases -- 138 and 141 cases on 29 and 30 March respectively and 108 cases (42 cases in the state quarantine) on 8 April 2020 -- may be linked to a southern Muslim pilgrimage defying health advisories and took part in the trip amid the outbreak. The official confirmed a spreading of the COVID-19 virus during the religious ceremony in Dawah, Indonesia. On 15 March 2020, the Thai Government started searching for 132 high COVID-19 infectious risk people returning from Dawah. This search was approximately 14 days before the surges on 29 and 30 March 2020. On 6 April 2020, a group of 42 Thais returned from Dawah, and a spike of 108 cases was documented on 8 April 2020.

In the Intervention II stage, policies to control the transmission route (nationwide curfew; banning of all international flights and mandatory state quarantine for all in-bound passengers) were introduced to reduce the frequency of contact and prevent SARS-CoV-2 from spreading to Thailand. At the end of the Intervention II stage, the number of daily confirmed cases was stable and very low. After Thailand had the number of daily confirmed cases below 10 cases for seven consecutive days, the Thai Government announced the Easing stage and began relaxing restrictions imposed during the Intervention I and II stages.

As a positive side-effect, the changes in human behavior (awareness in handwashing, facial mask-wearing, and social distancing) during the COVID-19 outbreak may contribute to the decreased transmission of other respiratory tract infections, such as influenza [9].

### Easing periods

After the number of daily confirmed cases was lower than 10 cases for seven consecutive days, the Thai Government started to relax the public health policies.

The Easing stage is from 3 May 2020 to 31 July 2020, where the restrictions corresponding to the lower infectious risk were lifted and observed for 14 days before proceeding to the next level, which eased the policies related to a higher infectious risk. The Easing stage was classified into five easing phases, and Thailand was in the fifth easing phase by the end of the first wave.

Fig 5 showed five easing phases. The first Easing phase had locally confirmed cases almost every day, and the highest number of cases was on 11 May 2020, which had 6 cases. The average of the locally confirmed cases in the first and second Easing phases were 1.57 and 0.6 cases, respectively. It can be seen that the locally confirmed cases declined to zero in the late of the second Easing phase. The number of locally confirmed cases was never reported again during the third, fourth, and fifth Easing phases.

**Fig 5.**
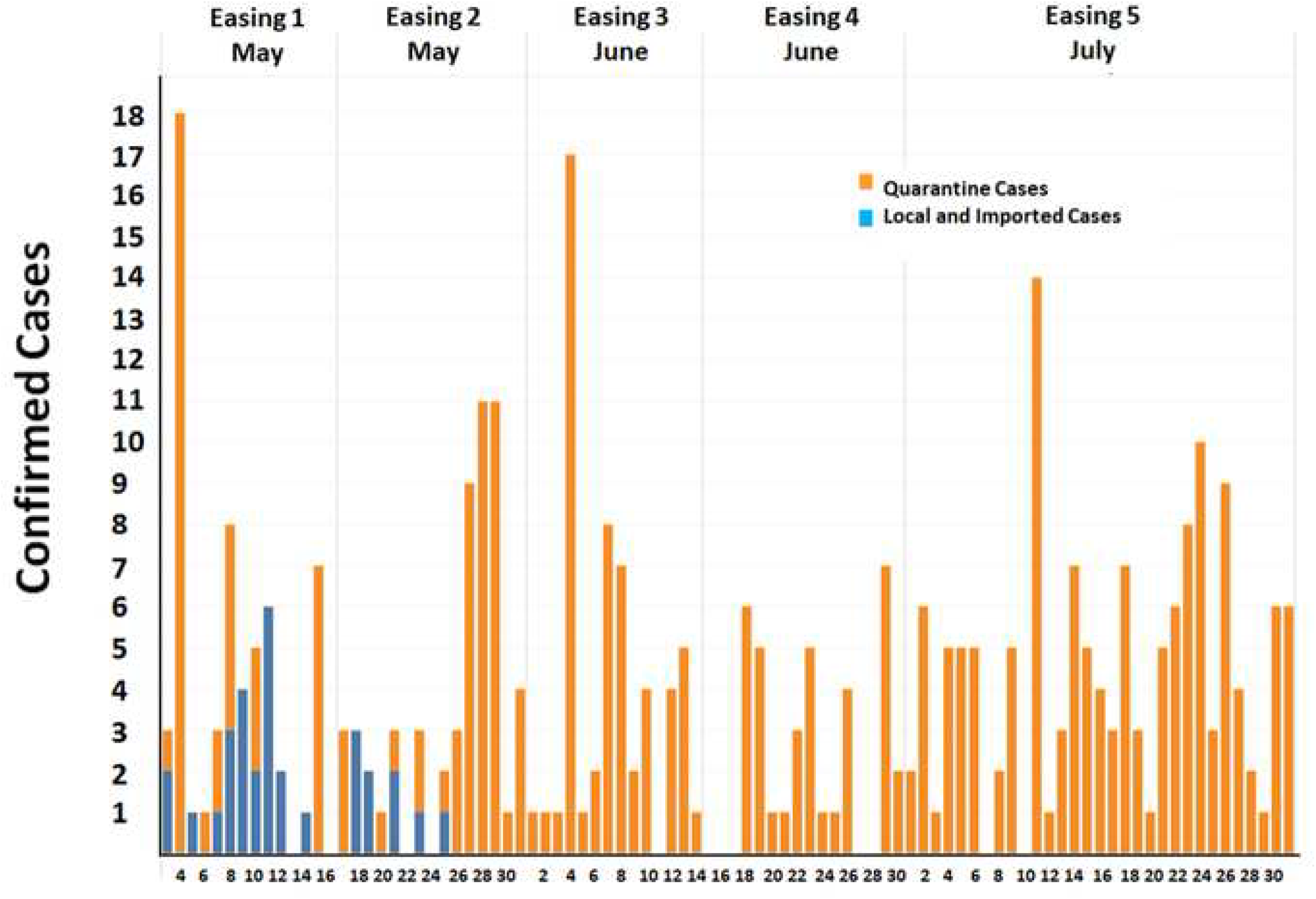
Thailand’s daily confirmed cases during the Easing stage (3 May to 31 July 2020).

Thai Government declared an easing state in five phases starting from 3 May to 31 July 2020 [16]. Each phase consisted of activities [16] shown in Table 2. The results showed that each easing phase had a different risk score, scale from 1 to 9, modified from the Texas Medical Association [17]. The average risk scores ranged from 3.44 to 7.75, depending on the activities in each phase. The Easing phase I had the lowest risk scores followed by phases II, III, IV, and V.

**Table 2.**
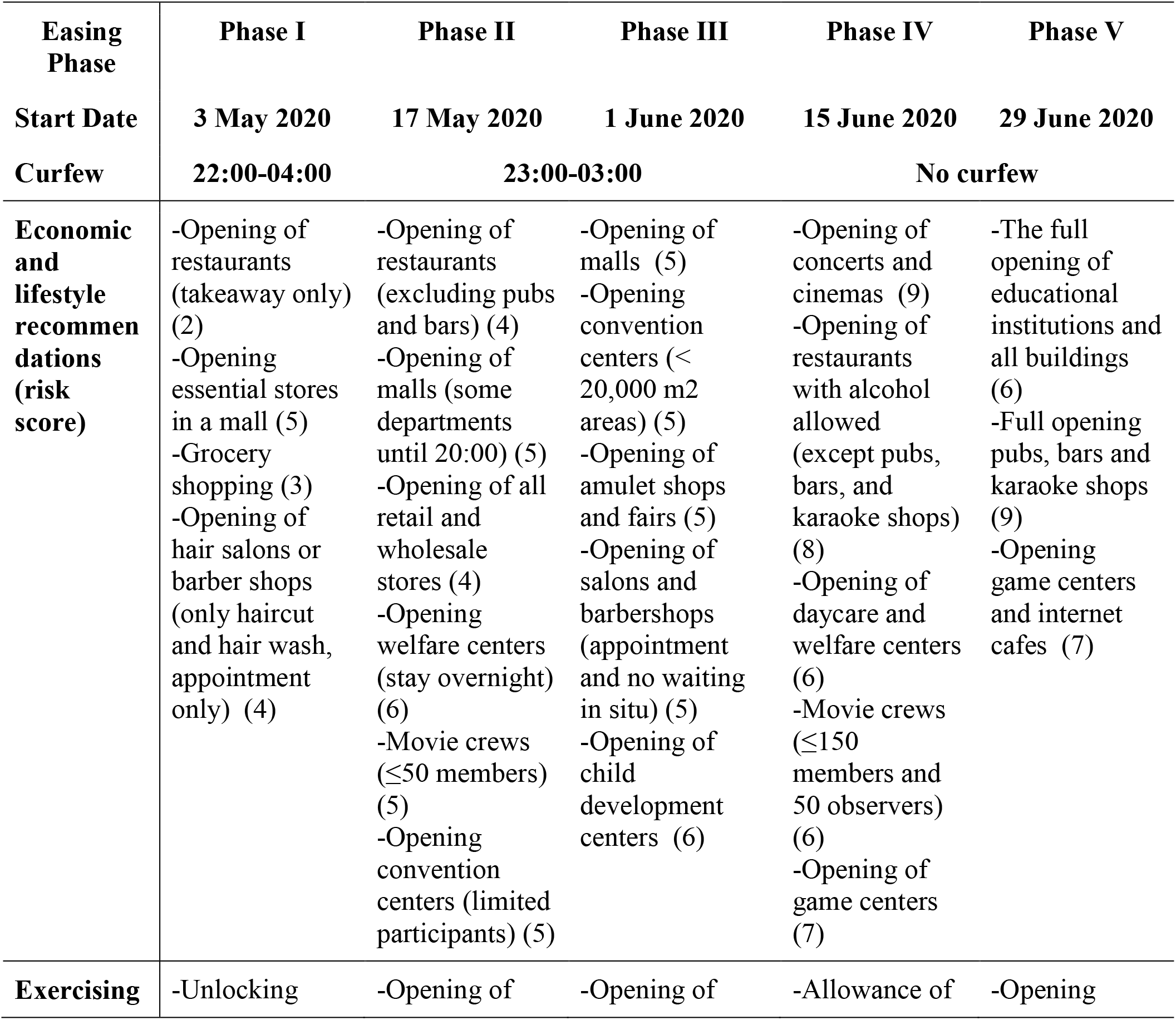
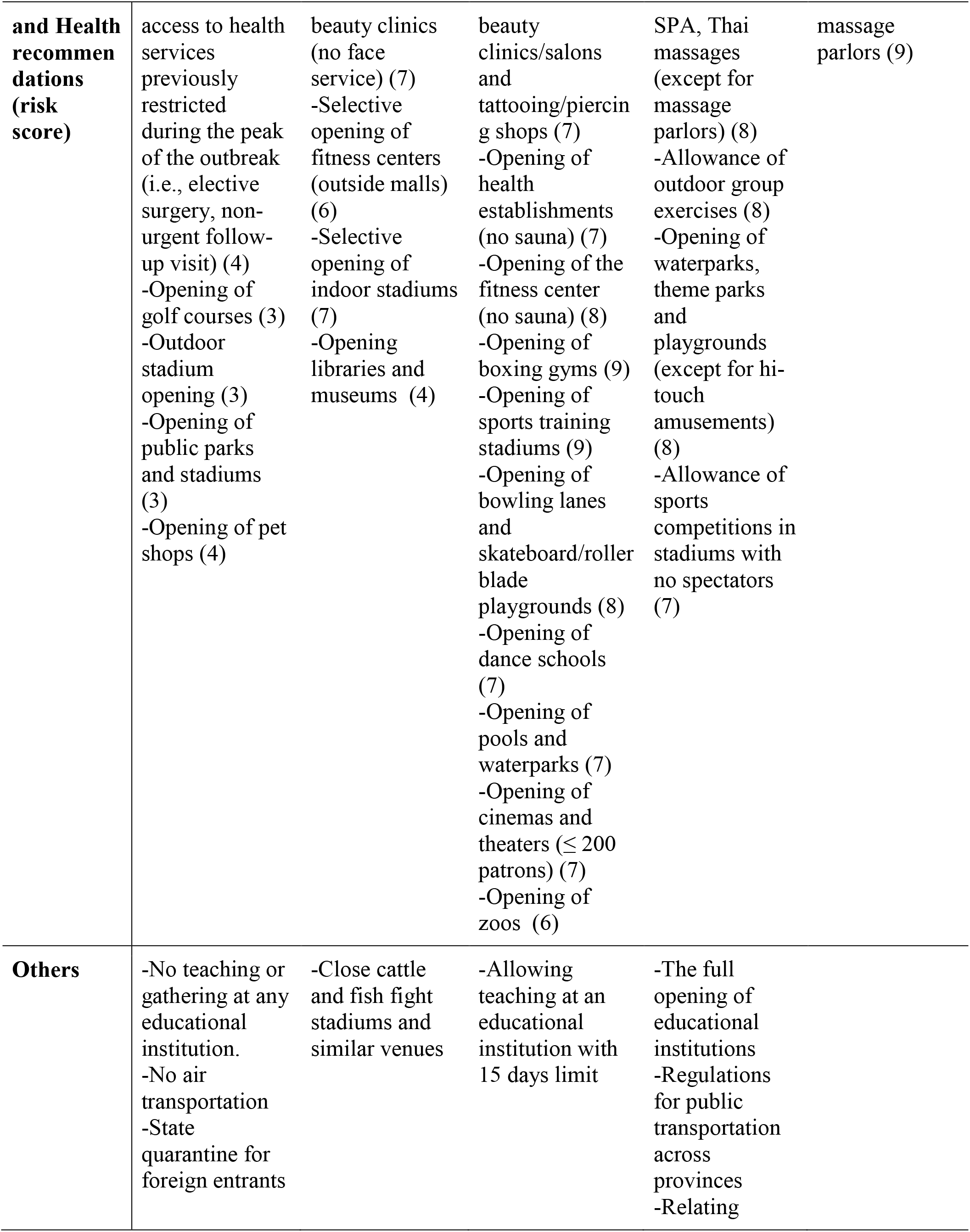
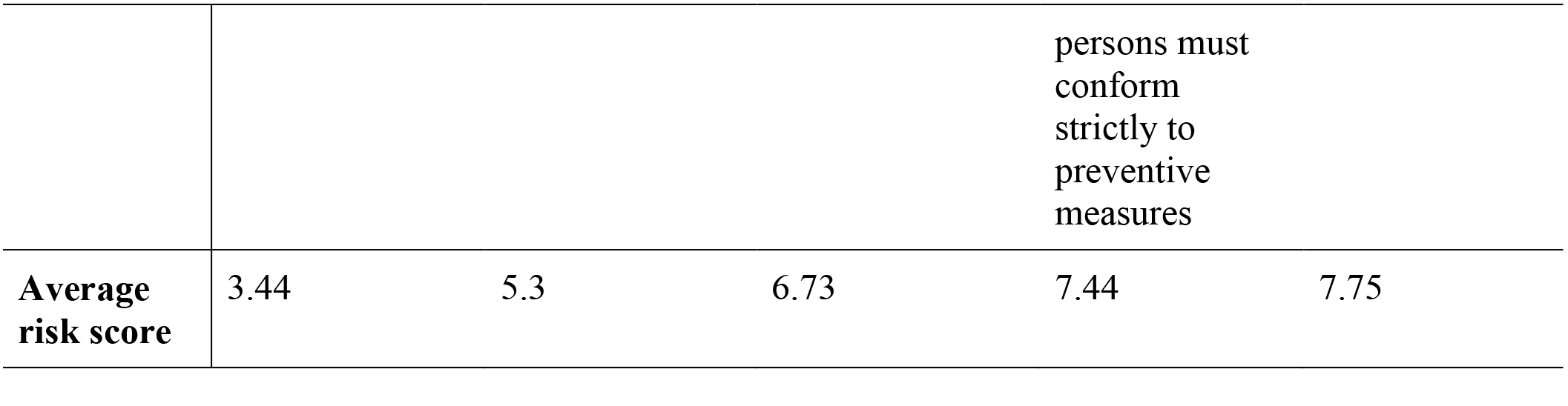
Easing phases, activities and risk scores for COVID-19 in Thailand.

### Economic Impacts

In retrospect of some previous studies comparing the effect of the different spectrum of intensity of interventions on different countries, it would seem that Thailand has chosen the right approach of adopting the most vigorous interventions as soon as possible to quickly control the COVID-19 epidemic before alleviating them after having the disease under control. However, at what cost Thailand paid for it, as far as the economy is concerned. Our discussion here meant to scratch the surface on this question.

To understand the trade-off of the two different COVID-19 handling approaches on health and economics, we select two neighbors in Scandinavian, Sweden, and Denmark, as subjects of our analysis. On the one hand, Sweden chose to intervene lightly to avoid severe impacts on the economy and believe that eventually, the herd immunity will kick in and stop the spreading. There was almost no lockdown nor closure of business, restaurants, and bars in Sweden. On the other hand, Denmark adopted all necessary measures to contain the outbreak at all costs. Almost all businesses and other places are closed down, and social distancing is applied wherever possible.

After all, we observe that these two approaches yield a similar projection on the Gross Domestic Product (GDP) as shown in Table 3. Although Sweden’s economics seem to be slightly better than Denmark in the first quarter of 2020, 0.1 percent versus −2.1 percent, respectively, the International Money Fund (IMF) predicts the GDP of Sweden and Denmark for the entire 2020 as −6.8 and −6.5 percent respectively. In addition, IMF predicts the unemployment rate, another key measure on economics, of Sweden and Denmark for the entire 2020 as 10.1 (increased from 6.8% in 2019) and 6.5 (increased from 5% in 2019) percent, respectively.

**Table 3.** Gross Domestic Product and unemployment rate in Sweden and Denmark.

| Country | Gross Domestic Product (%) |  |  |  |  |  | Unemployments (%)** |  |  |
| --- | --- | --- | --- | --- | --- | --- | --- | --- | --- |
|  | Present |  | Projections |  |  |  | Present | Projections |  |
|  | 2019 | 2020Q1<br>* | 2020Q2<br>* | 2020Q3<br>* | 2020Q4<br>* | 2020 | 2019 | 2020 | 2021 |
| **Sweden** | 1.2 | 0.1 | -9.1 | 1.9 | 1.7 | -6.8 | 6.8 | 10.1 | 8.9 |
| **Denmark** | 2.4 | -2.1 | -8.6 | 3.9 | 3.0 | -6.5 | 5.0 | 6.5 | 6.0 |
| **Thailand** | 2.4 | N/A | N/A | N/A | N/A | -6.7 | 1.1 | 1.1 | 1.1 |

Sources: IMF[18], \*Europa[19]
\*\* National definitions of unemployment may differ.

However, as reported in Table 4 the impacts on public health of the two approaches are enormous. As of 31 July 2020, the total number of confirmed COVID-19 infected cases in Sweden and Denmark are 7,931.27 and 2,369.57 cases per million populations (more than 3.3 times difference). The total number of deaths due to coronavirus in Sweden is 568.26 deaths per million population, whereas the death in Denmark on the same day is 106.18 deaths per million. Essentially, Sweden has more than five times higher mortality rate than Denmark. In the short run, the strict public intervention policies seem to be in favor; however, the impacts, in the long run, will remain to be seen.

**Table 4.** Infected cases and deaths caused by COVID-19 in Sweden, Denmark and Thailand on 31 July 2020.

| Country | Infected cases* (per million people) | Deaths** (per million people) |
| --- | --- | --- |
| Sweden | 7931.27 | 568.26 |
| Denmark | 2369.57 | 106.18 |
| Thailand | 47.42 | 0.83 |

Sources: \*Our world in data: Total confirmed COVID-19 cases per million people [21]
\*\*Our world in data: Total confirmed COVID-19 deaths per million people [20]

Thailand also strictly practiced social distancing and lockdown, and the IMF predicts the GDP of the entire 2020 to −6.7 percent. As of 31 July 2020, the total number of confirmed COVID-19 infected cases in Thailand is 47.42 cases per million populations. Moreover, the total confirmed COVID-19 deaths on 31 July 2020 is 0.83 deaths per million people, which is among the world’s lowest mortality rates. The interesting question is, while Thailand seems to be doing very well with the lockdown policy, how the global pandemic situation affects some key sectors, such as tourism, which is among the main contributions to Thailand’s GDP, remains to be answered, especially if the second wave of COVID-19 hit Thailand.

In essence, thus far, both approaches to handling the COVID-19 outbreak seem to bear similar impacts on the economy; however, adopting more potent public health interventions leads to more survival. Please note that even though we use two key economic indicators (GDP and unemployment rate) to quantify the impacts on the economy briefly, our discussion is not to replace a thorough investigation of the economic impact.

## Conclusions

Among the top destinations of tourists from Wuhan is Thailand. Thailand was among the first countries outside China to find COVID-19 cases. Nevertheless, Thailand exited the first wave of the COVID-19 outbreak in less than six months, with 3,072 recoveries (about 96.15%) out of 3,195 infectious cases, including stage quarantine cases.

We detailed how Thais counteracted the threat of the COVID-19 pandemic and their sacrifices. Apart from the two super-spreading events, as Bangkok was shut down without a national-level plan to handle outward mobilization, a massive migration of labor force back to their hometowns was triggered, spreading COVID-19 nationwide. Nevertheless, at an early stage of the pandemic, Thais responded cooperatively with personal-hygiene and protection guidelines. In the later stage of the pandemic, Thais cooperated enthusiastically with physical distancing and policies disallowing social gatherings. Most of the public and private sectors transitioned to a work-from-home environment. These norms helped to reduce the chances of infection considerably.

The Thai Government had a significant role in the initial stage of the crisis. In particular, it established the CCSA to oversee overall problems holistically, facilitate the integration of all related bodies, and ensure consistent and unambiguous communications. Despite unavoidable mistakes (such as its inability to stop the massive migration that happened two weeks after its inception), the CCSA learned from its mistakes, adapted, improved, and became more effective afterward. The Thai Government will continue to have a significant role in managing the pandemic.

In terms of observation on the economic impact, trying to intervene slightly can slow down the GDP to decline momentarily. Using herd immunity or lockdown approaches to handle the COVID-19 outbreak seem to bear similar impacts on the economy; however, adopting more potent public health interventions leads to more survival.

If the second wave of the COVID-19 outbreak hits Thailand, the Thai Government needs practical tools for preventive policy planning. These tools should be data-driven, in real-time, and aid decision-making rapidly. We used some of these data-driven tools to form visual and measurement analytics (Fig 3, Fig 4, and Table 2). We plan to utilize these tools or develop new data-driven tools with good predictive power to help tackle a second outbreak of COVID-19 in Thailand.

## Data Availability

The study in this article used the medical records of laboratory-confirmed COVID-19 patients in Thailand from 12 January 2020 to 31 July 2020. These medical records were retrieved from the Department of Disease Control of Thailand website given below.

https://ddc.moph.go.th/viralpneumonia/eng/index.php

